# Cardiometabolic Risk Trajectories in Young Adults Across 16 Middle East and North African Countries, 1990–2018: A Sex-Asymmetric Divergence

**DOI:** 10.64898/2026.09.12.26362914

**Authors:** Marwan Kobtan, Shatha Al Aqarbeh, Yara Shhab, Nada Soliman, Shahinazhanem Abdellatif, Adalia Cristiana Iancu, Shady Elrashidy, Mustafa Al Majali

## Abstract

**Background:** The INTERHEART–Middle East study identified the Middle East and North Africa (MENA) region as having the youngest age at first acute myocardial infarction (AMI) globally, with a population-attributable risk of 97.5% across nine modifiable risk factors. The population-level trajectories of cardiometabolic risk factors that precede this premature MI signature in young MENA adults remain incompletely characterized.

**Methods:** NCD Risk Factor Collaboration (NCD-RisC) modeled prevalence estimates were analyzed across 16 MENA countries (Bahrain, Egypt, Iran, Iraq, Jordan, Kuwait, Lebanon, Libya, Morocco, Oman, Qatar, Saudi Arabia, Syria, Tunisia, United Arab Emirates, Yemen) in adults aged 20–29 and 30–39 years, stratified by sex: obesity and diabetes prevalence through 2022, hypertension prevalence through 2019 (ages 30–39 only, by data availability), and non-HDL cholesterol through 2018. Five-year age bands were aggregated into 20–29 and 30–39 groups by equal-weighted mean (with UN World Population Prospects 2024 population-weighted aggregation tested as a sensitivity analysis). Each country × sex × age-band × riskfactor series was analyzed using (1) joinpoint regression for the average annual percent change (AAPC); (2) posterior probability of true 1990-to-endpoint change derived from published credible intervals; and (3) a mixed-effects model with a year × region (MENA vs. rest-of-world [ROW]) interaction and country-level random slopes. Sensitivity analyses excluded conflict-affected states and Iran.

**Results:** MENA diverged significantly from ROW in 6 of 14 sex × age-band comparisons, with a sex-asymmetric direction. In men, MENA worsened faster than ROW: obesity, ages 20–29 (AAPC +3.49%/year; p=0.009); obesity, ages 30–39 (AAPC +2.65%/year; p=0.043); and diabetes, ages 30–39 (AAPC +1.90%/year; p=0.014). In women, MENA improved faster than ROW: hypertension, ages 30–39 (AAPC −0.58%/year; p=0.001); non-HDL cholesterol, ages 20–29 (p=0.017); and non-HDL cholesterol, ages 30–39 (p=0.017). Country-level posteriors confirmed near-universal regional patterns: 60 of 64 obesity cells showed a very probable increase since 1990, and 52 of 64 (81%) non-HDL cholesterol cells showed a probable or very probable decrease. Five of the six divergent findings survived correction for multiple testing.

**Conclusion:** Cardiometabolic risk in young MENA adults is diverging from global patterns in a sex-asymmetric way: obesity and diabetes are accelerating in young men, while hypertension and lipid profiles are improving in young women. This pattern maps directly onto the young-male AMI signature reported by INTERHEART–Middle East and identifies obesity and diabetes in young men as the most urgent targets for regional primary prevention.

## Introduction

Cardiovascular disease is the leading cause of death in the Middle East and North Africa (MENA), and its clinical onset in this region occurs earlier in life than in any other world region studied to date. The INTERHEART Middle East study reported a mean age at first AMI of 51.2 ± 10.3 years — the youngest of any region in the INTERHEART program — with the largest proportion of patients presenting before age 40 [1]. Nine modifiable risk factors accounted for a population-attributable risk (PAR) of 97.5% for AMI in the region [1]. These findings established that the excess and premature cardiovascular burden in MENA reflects a markedly higher regional concentration of modifiable cardiometabolic risk rather than unmeasured or nonmodifiable susceptibility.

Despite this well-established clinical observation, no published analysis has characterized the population-level trajectory of cardiometabolic risk factors specifically within the 20–39-year age band across MENA. Global epidemiological surveillance, including the standard outputs of the NCD Risk Factor Collaboration (NCD-RisC), reports risk-factor estimates for all adults aged 18 years and older or restricts age-specific modeling to the 30–79-year band; hypertension, in particular, is not modeled below age 30 in the most recent global release. As a result, the early-adulthood window in which atherosclerotic and metabolic pathology first initiates remain uncharacterized in regional surveillance, and young MENA adults are effectively invisible in the existing literature linking population-level risk-factor trends to the region’s premature coronary disease burden.

The objective of this study was to characterize 30-year secular trends (1990–2018) in four cardiometabolic risk factors — obesity, diabetes, hypertension, and non-HDL cholesterol — among adults aged 20–29 and 30–39 years across 16 MENA countries, and to formally test whether these trends differ from those observed across 184 non-MENA “rest-of-world” (ROW) countries over the same period. Trends were quantified using joinpoint regression to estimate the average annual percent change (AAPC), Bayesian posterior probability modeling to classify the direction and certainty of change since 1990, and linear mixed-effects models with a year × region interaction term to formally test divergence between MENA and ROW trajectories, stratified by sex and age band.

## Methods

### Data Sources & Harmonization

Country-level risk-factor estimates were obtained from the NCD Risk Factor Collaboration (NCD-RisC; ncdrisc.org), a global research consortium that produces Bayesian hierarchical model estimates by pooling individual-level data from population-representative health surveys worldwide. Four risk factors were drawn from the most recent NCD-RisC release available for each: diabetes prevalence (data through 2022) [2], body-mass index (BMI) and obesity prevalence (data through 2024) [3], hypertension prevalence (data through 2019, ages 30 years and older only) [4], and non-HDL cholesterol concentration (data through 2018) [5]. Because the non-HDL cholesterol release sets the latest year common to all four risk factors, the primary analytic window for this study was fixed at 1990–2018 across all outcomes, notwithstanding the longer follow-up available in the underlying diabetes and BMI/obesity source files. As all four outcomes are modeled estimates rather than raw survey measurements, the uncertainty inherent to the underlying Bayesian hierarchical models was not propagated into the downstream trend statistics described below (Tiers 1 and 3); this uncertainty was incorporated separately in Tier 2, via the published credible intervals.

NCD-RisC publishes diabetes, BMI/obesity, and hypertension estimates in 5-year age bands (20–24, 25–29, 30–34, 35–39, …); the non-HDL cholesterol release is structured directly in 10-year bands beginning at 20–29 and required no aggregation. For the remaining three risk factors, the two component 5-year bands were combined into 20–29 and 30–39 groups by an equal-weighted mean across country-year-sex strata in the primary analysis. As a pre-specified sensitivity analysis, this aggregation step was repeated using UN World Population Prospects (WPP) 2024 mid-year population counts as weights — prevalence_20-29_ = (pop_20-24_× prev_20-24_ + pop_25-29_ × prev_25-29_) / (pop_20-24_+ pop_25-29_) — to test whether population-weighting the two component bands materially altered the aggregated estimate (see Sensitivity Analyses, below).

### Geographical Framework & Outcomes

Sixteen MENA countries were pre-specified before data extraction: Bahrain, Egypt, Iran, Iraq, Jordan, Kuwait, Lebanon, Libya, Morocco, Oman, Qatar, Saudi Arabia, Syria, Tunisia, the United Arab Emirates, and Yemen. These were compared against the remaining 184 non-MENA countries present in the NCD-RisC files, pooled as a single “rest-of-world” (ROW) comparator. For consistency, the ROW trajectory used throughout is the population-weighted mean across these 184 non-MENA countries, not the NCD-RisC-published “World” aggregate, which itself includes the 16 MENA countries and would otherwise contaminate the region contrast.

Four cardiometabolic outcomes were extracted, each stratified by sex and by age band (20–29 and 30–39 years):

- **Obesity** — prevalence of body-mass index ≥30 kg/m^2^
- **Diabetes** — prevalence of diabetes, defined as fasting plasma glucose ≥7.0 mmol/L, glycated hemoglobin (HbA1c) ≥6.5%, or use of antidiabetic medication
- **Hypertension** — prevalence of systolic blood pressure ≥140 mmHg, diastolic blood pressure ≥90 mmHg, or use of antihypertensive treatment; restricted to the 30–39-year age band, as the source release does not model blood pressure below age 30
- **Non-HDL cholesterol** — mean non-HDL cholesterol concentration, mmol/L Because hypertension is not estimable for the 20–29-year band, the full 4-risk-factor × 2-sex × 2-age-band matrix yielded 14 analyzable cells rather than 16 (the two 20–29 hypertension cells were excluded).

### Statistical Analysis: Three Analytical Tiers

All statistical analyses were performed in R, applied independently to each country × sex × ageband × risk-factor series.

#### Tier 1 — Joinpoint regression for average annual percent change (AAPC)

For each series, a piecewise log-linear regression of the annual estimate on calendar year was fit using the segmented R package. Candidate models with 0, 1, and 2 breakpoints were compared, and the best-fitting model was selected by Bayesian Information Criterion (BIC). The annual percent change (APC) for each segment was computed as (exp(β) − 1) × 100, where β is the segment slope on the log scale; the AAPC across 1990–2018 was calculated as the segment-length-weighted average of segment APCs, with 95% confidence intervals derived via the delta method. This tier generated 224 country-level trajectories in total (16 countries × 14 sex/age-band/riskfactor cells)

#### Tier 2 — Bayesian posterior probability of true change

For each country × cell, the posterior probability that the 2018 value exceeded the corresponding 1990 value was estimated from the published NCD-RisC 95% uncertainty intervals (UI), assuming an approximately normal posterior on the log scale with standard deviation σ = (log [UI upper] − log [UI lower]) / (2 × 1.96) at each time point. The log-difference between 2018 and 1990 was treated as normally distributed, with mean equal to the difference in log point estimates and variance equal to the sum of the two time-point variances; P (2018 > 1990) was derived from this distribution.Countries were classified as: very probable increase (P > 0.95), probable increase (0.80 < P ≤ 0.95), uncertain (0.20 ≤ P ≤ 0.80), probable decrease (0.05 ≤ P < 0.20), and very probable decrease (P < 0.05) — mirroring NCD-RisC’s own directional-reporting convention.

#### Tier 3 — Linear mixed-effects model, MENA vs. rest-of-world

For each risk factor × sex × age band, a linear mixed-effects model was fit as value ~ year_centered * region + (year_centered | country), where *region* is a binary factor (MENA vs. ROW), *year_centered* is calendar year centered at the study midpoint (2004), and *country* carries a random intercept and random slope on year. Models were fit by restricted maximum likelihood (lme4: lmer), with fixed-effect p-values derived from Satterthwaite-approximated degrees of freedom (lmerTest). The year_centered × region interaction coefficient — the annual difference in rate of change between MENA and ROW — was the pre-specified headline statistic for each of the 14 outcome cells

### Sensitivity Analyses

Five pre-specified sensitivity analyses were conducted:

1. **UN WPP 2024 population-weighted aggregation** — the 5-year-to-10-year age-band aggregation step (see Data Sources & Harmonization) was repeated using WPP 2024 population counts as weights, in place of the equal-weighted mean used in the primary analysis.
2. **Exclusion of conflict-affected states** — Syria, Yemen, Libya, and Iraq were removed from the MENA group (12 of 16 countries retained), to assess whether estimates from countries with markedly reduced survey coverage during periods of conflict were driving the regional signal.
3. **Exclusion of Iran** — Iran, sometimes analyzed separately in MENA cardiovascular epidemiology given its population size and distinct risk-factor profile, was removed (15 of 16 countries retained).
4. **Gulf Cooperation Council (GCC)-only subgroup** — the MENA group was restricted to the 6 GCC countries, to test whether findings held within this more economically and epidemiologically homogeneous subgroup.
5. **Multiple-comparisons correction** — Benjamini-Hochberg false discovery rate (FDR) correction was applied to the 14 primary Tier 3 interaction p-values, with statistical significance redefined as q < 0.05.

### Limitations

Four limitations are acknowledged. First, all risk-factor estimates are NCD-RisC Bayesian hierarchical model outputs pooling multiple population-representative surveys, not raw survey measurements; Tiers 1 and 3 treat these modeled point estimates as observed values, with modeled-estimate uncertainty incorporated only in Tier 2, via the published credible intervals.

Second, hypertension data are restricted to ages 30 years and older in the current NCD-RisC release; the 20–29 hypertension trajectory could not be quantified and is not reported. Third, the non-HDL cholesterol endpoint is 2018, the most recent year available in the current release, which set the common analytic endpoint for all four risk factors; trends beyond 2018 cannot be assessed until an updated cholesterol release is published. Fourth, country-year uncertainty intervals are wider where survey coverage is sparse, notably in Syria, Yemen, and Libya after 2011, and the ecological, country-level design of this analysis does not support individual-level causal inference.

## Results

### 1. Primary Regional Divergence: A Sex-Asymmetric Pattern

Of the 14 analyzable risk-factor × sex × age-band cells, 6 (43%) showed MENA diverging significantly from ROW on the Tier 3 mixed-effects year × region interaction. The direction of divergence was strictly sex-asymmetric: all 3 significant worsening trajectories occurred in young men (obesity at both age bands, diabetes at 30–39 years), and all 3 significant improving trajectories occurred in young women (hypertension at 30–39 years, non-HDL cholesterol at both age bands). No risk factor showed a significant divergence of the same direction in both sexes.

### 2. Accelerated Cardiometabolic Risk in Young Men

In men, MENA prevalence rose significantly faster than ROW for three of the seven male cells assessed:

- **Obesity, ages 20–29:** AAPC +3.49%/year (country range +1.60 to +5.21); Δslope vs. ROW +0.13 percentage points (pp)/year; p = 0.009.
- **Obesity, ages 30–39:** AAPC +2.65%/year (range +1.29 to +4.24); Δslope +0.11 pp/year; p = 0.043.
- **Diabetes, ages 30–39:** AAPC +1.90%/year (range −0.76 to +4.38); Δslope +0.07 pp/year; p = 0.014.

### 3. Favorable Trends in Young Women

In women, MENA prevalence or concentration improved significantly faster than ROW for three cells:

- **Hypertension, ages 30–39:** AAPC −0.58%/year (country range −3.40 to +1.06); Δslope vs. ROW −0.15 pp/year; p = 0.001.
- **Non-HDL cholesterol, ages 20–29:** AAPC −0.36%/year (range −0.92 to +0.18); Δslope −0.0079 mmol/L/year; p = 0.017.
- **Non-HDL cholesterol, ages 30–39:** AAPC −0.38%/year (range −0.99 to +0.19); Δslope −0.0091 mmol/L/year; p = 0.017.

The remaining 8 cells showed no significant MENA-ROW divergence. Obesity in women rose in absolute terms at both age bands but did not differ significantly from the ROW trajectory (Δslope slightly negative, non-significant). Diabetes in men aged 20–29 and in women at both age bands rose in a pattern statistically consistent with the global trend. Hypertension in men aged 30–39 was essentially flat and did not diverge from ROW. Non-HDL cholesterol in men declined at both age bands but not differently from the ROW decline.

### 4. Posterior Probabilities and National Standouts

Country-level Bayesian posterior probabilities of a true 1990-to-2018 increase or decrease corroborated the direction of the Tier 3 findings and showed these patterns to be near-universal across the region rather than driven by a small number of countries:

- **Obesity** (4 cells × 16 countries = 64 country-cells): 60/64 classified as very probable increase; 4 uncertain; none showed a probable or very probable decrease.
- **Non-HDL cholesterol** (64 country-cells): 52/64 (81%) classified as probable or very probable decrease (32 very probable decrease, 20 probable decrease) — a near-universal regional decline.
- **Diabetes** (64 country-cells): 44/64 classified as probable or very probable increase (10 very probable, 34 probable); 20 uncertain; none showed a decrease.
- **Hypertension** (2 cells × 16 = 32 country-cells): 28/32 were uncertain at the country level; only 3/32 showed a probable decline, concentrated in higher-population countries (notably Iran and Tunisia), which pulled the regional pooled estimate toward decline.

Several countries were consistent outliers driving the regional signals. **Egypt** showed the region’s most extreme diabetes rise (men, ages 30–39: AAPC +4.38%/year). **Iran** showed both the strongest obesity rise among men (ages 20–29: AAPC +5.12%/year) and, independently, the strongest hypertension decline of any country in any of the six significant findings (women, ages 30–39: AAPC −3.40%/year). **Yemen** showed the region’s highest obesity rise (men, ages 20–29: AAPC +5.21%/year) and was the only MENA country with a rising, rather than falling, non-HDL cholesterol trajectory in women (ages 20–29: +0.18%/year; ages 30–39: +0.19%/year).

**Saudi Arabia** was the only MENA country with a declining diabetes trajectory in men at either age band (ages 30–39: AAPC −0.76%/year; ages 20–29: −1.27%/year). Tunisia and Morocco also showed prominent male obesity increases (Tunisia +5.01%/year, Morocco +4.11%/year, both ages 20–29), and Libya and Iraq were substantial contributors to the regional male diabetes signal (AAPC +3.41%/year and +2.74%/year, respectively, ages 30–39), while Qatar showed the region’s largest-magnitude non-HDL cholesterol decline in women (ages 20–29: −0.92%/year; ages 30–39: −0.98%/year).

### 5. Joinpoint Breakpoint Cascade

Country-level joinpoint regression (Tier 1) identified at least one statistically significant inflection point (Davies test, p < 0.05) in 220 of the 224 country-level trajectories. The mean breakpoint year followed a sequential cascade across risk factors and sexes:

1. Non-HDL cholesterol, both sexes and age bands — mean breakpoint ~2001 (range of means 2000.9–2001.3), the earliest to inflect.
2. Diabetes, women — mean breakpoint ~2002 (range 2002.2–2002.6).
3. Hypertension, women — mean breakpoint ~2004.
4. Hypertension, men — mean breakpoint ~2006.
5. Diabetes, men — mean breakpoint ~2007 (range 2006.9–2007.5).
6. Obesity, women — mean breakpoint ~2008 (range 2008.2–2008.3).
7. Obesity, men — mean breakpoint ~2009 (range 2009.3–2009.4), the latest to inflect.

The direction and statistical significance of segment slopes were robust across this cascade. The ordering is consistent with a sequential cardiometabolic transition in which lipid trajectories stabilized first, glycemic trends in women shifted next, blood pressure trends shifted mid-period, glycemic trends in men shifted later, and body-mass index shifted last.

### 6. Sensitivity Robustness

Applying Benjamini-Hochberg false discovery rate (FDR) correction to the 14 primary Tier 3 interaction p-values, 5 of the 6 significant findings survived at q < 0.05; only obesity in men aged 30–39 fell below this threshold (q = 0.101).

Country-exclusion sensitivity analyses clarified which countries were driving the two strongest single-country-influenced signals. For the hypertension decline in women aged 30–39, excluding Iran attenuated the pooled AAPC from −0.58%/year to −0.39%/year (a 32% reduction in magnitude) but the interaction remained highly significant (p = 0.006 without Iran), indicating that Iran amplifies, but does not solely create, the regional improvement signal. For the diabetes rise in men aged 30–39, the signal was materially different under two exclusion sensitivities: restricting to the 6 GCC countries removed the significant interaction entirely (p = 0.681), and excluding the four conflict-affected states (Syria, Yemen, Libya, Iraq; Egypt retained) also attenuated it substantially (p = 0.162). Because Egypt (AAPC +4.38%/year) remained in both of these subsets while the signal nonetheless weakened or disappeared, the diabetes rise in young men is attributable to a cluster of non-GCC countries collectively — Egypt, Libya (+3.41%/year), Iraq (+2.74%/year), Iran (+2.53%/year), and Yemen (+2.35%/year) — rather than to any single country. Conversely, the GCC-only subgroup confirmed that the male obesity signal at ages 20–29 was not an artifact of non-GCC countries: the interaction remained significant, and numerically stronger, within the GCC alone (p = 0.002), while the female hypertension improvement at ages 30–39 also persisted within the GCC subgroup, albeit attenuated by the smaller sample (p = 0.027).

## Discussion

### Principal Findings

In this analysis of 16 MENA countries over three decades, young adults in the region are undergoing a sex-divergent cardiometabolic transition rather than a uniform one. Among men, obesity and diabetes accelerated significantly faster than the rest of the world across early and later young adulthood; among women, hypertension and non-HDL cholesterol improved significantly faster than the rest of the world over the same period. No risk factor diverged from the global pattern in the same direction in both sexes. This divergence was not confined to a small number of atypical countries: country-level posterior probabilities showed the male obesity rise and the female non-HDL cholesterol decline to be near-universal across the 16-country panel, and five of the six divergent findings survived correction for multiple comparisons.

### The INTERHEART–Middle East Bridge

These trajectories offer a population-level explanation for a clinical observation reported a decade earlier. The INTERHEART Middle East study established that the region’s patients present with first acute myocardial infarction (AMI) at the youngest mean age of any world region (51.2 ± 10.3 years), with the largest proportion of patients under 40, and that nine modifiable risk factors accounted for 97.5% of the population-attributable risk for AMI in the region [1]. The present findings map directly onto what has been described as the young-male AMI signature of the region: obesity and diabetes are precisely the factors accelerating fastest, and earliest, from ages 20–29 onward, in MENA men relative to the rest of the world. This suggests that the region’s young-male AMI signature is not a fixed epidemiological feature but the downstream consequence of a risk-factor trajectory that begins diverging from global patterns in early adulthood, well before the age at which cardiovascular screening conventionally begins.

### Sociocultural and Healthcare Drivers

This ecological analysis cannot directly adjudicate among candidate explanations for the sex-asymmetric pattern observed, but several mechanisms discussed elsewhere in the regional literature are plausible contributors. A regional nutrition transition — increasing availability and consumption of ultra-processed, energy-dense foods alongside rapid urbanization — has been proposed as a driver of rising obesity prevalence across MENA generally; if this transition has disproportionately affected dietary intake and occupational activity among young men, including a shift toward more sedentary employment, it could plausibly help explain a male-concentrated obesity and diabetes rise. Differential healthcare engagement between the sexes offers a complementary, non-exclusive explanation for the female-favorable trends: young women in many MENA health systems have recurring contact with clinical services through maternal and reproductive care, creating incidental opportunities for blood pressure and lipid screening and counseling, whereas young men in the same health systems typically have little contact with a physician before middle age. Neither mechanism was measured directly in this study, and the relative contribution of dietary, occupational, and health-system factors to the sex asymmetry reported here should be regarded as hypothesis-generating rather than established.

### National Heterogeneity

MENA is not a monolithic risk environment, and the country-exclusion sensitivity analyses indicate that the region’s two worsening male signals have different underlying architectures. The diabetes rise in men aged 30–39 was not attributable to any single country: Egypt showed the highest country-level AAPC (+4.38%/year), but Libya (+3.41%/year), Iraq (+2.74%/year), Iran (+2.53%/year), and Yemen (+2.35%/year) each contributed substantially, and the MENAROW interaction disappeared entirely when the analysis was restricted to the six high-income Gulf Cooperation Council (GCC) countries (p = 0.681) and attenuated substantially when the four conflict-affected states were excluded (p = 0.162). This pattern is consistent with a diabetes signal concentrated among non-GCC countries with more constrained health-system resources, including states with substantial recent conflict-related disruption, although this ecological analysis cannot establish conflict exposure as a direct cause. By contrast, both the male obesity rise at ages 20–29 and the female hypertension improvement at ages 30–39 persisted within the GCC subgroup alone (p = 0.002 and p = 0.027, respectively), indicating that these two signals are region-wide phenomena and not artifacts of the lower-income, non-GCC countries in the panel. Iran was the single largest contributor to the hypertension improvement in women (AAPC −3.40%/year, the largest-magnitude finding among all six divergent cells), yet the regional signal persisted, only partially attenuated, when Iran was excluded (AAPC −0.39%/year; p = 0.006) — indicating that Iran amplifies, rather than solely creates, this improvement.

### Clinical and Policy Implications

These findings carry a direct implication for cardiovascular prevention practice in MENA. Current cardiovascular risk-screening guidelines, including most regional adaptations of international lipid and diabetes screening recommendations, do not typically recommend routine cardiometabolic screening before age 40. The data presented here show that obesity and diabetes prevalence in MENA men are already diverging from global trends by ages 20–29 — two decades before that conventional threshold — in a population already known to present with first myocardial infarction earlier than in any other world region. Aligning cardiometabolic screening in MENA men — at minimum, body-mass index and fasting glucose or HbA1c — with the age at which regional risk-factor divergence actually begins, rather than with a threshold derived from later-onset disease patterns elsewhere, represents a concrete, actionable target for regional primary-prevention policy.

## Strengths and Limitations

This study’s principal strength is its formal statistical framework: rather than describing trends, it tests them, using three complementary methods (joinpoint AAPC, Bayesian posterior probability, and a mixed-effects region interaction) across the largest available country comparator panel, and confirms the primary findings against five pre-specified sensitivity analyses and multiplicity correction. Several limitations, already noted in the Methods, bear directly on interpretation. First, this is an ecological, country-level analysis; it identifies population-level divergence, not individual-level risk or causation, and cannot establish that the individuals gaining weight or developing diabetes are the same individuals who will experience premature myocardial infarction. Second, all outcome data are NCD-RisC Bayesian hierarchical model estimates pooling heterogeneous underlying surveys, not raw measurements, and the joinpoint and mixed-effects analyses treat these modeled point estimates as observed values.

Third, hypertension could not be assessed at ages 20–29 in the current global NCD-RisC release, so the earliest hypertension trajectory in young MENA adults remains uncharacterized; if a divergence analogous to the 30–39 female pattern exists at 20–29, it is not captured here. Fourth, the non-HDL cholesterol endpoint of 2018 constrained the entire analytic window; more recent lipid trends, including any response to the accelerating male obesity and diabetes signals described here, cannot yet be assessed.

## Conclusion

Cardiometabolic risk in young MENA adults is diverging from global patterns along sex-specific lines: obesity and diabetes are accelerating in men from as early as age 20, while blood pressure and lipid profiles are improving in women. This divergence offers a population-level explanation for the region’s already well-documented burden of premature myocardial infarction, which disproportionately affects men at the youngest mean age of any world region. Extending cardiometabolic screening in MENA men to early adulthood, rather than waiting until age 40, is a concrete step toward primary prevention that these findings directly support.

## Declarations

### Data Availability

All data are publicly available from the NCD Risk Factor Collaboration (NCD-RisC; ncdrisc.org).

### Declaration of Generative AI in Scientific Writing

During the preparation of this work, the authors used Claude 5 Sonnet (Anthropic) strictly for language editing and structural formatting of the manuscript prose.

After using this tool, the authors conducted an exhaustive line-by-line audit, reviewed and edited the content as needed, and take full responsibility for the absolute accuracy and integrity of the final publication. No AI was used in data collection, statistical analysis, or the generation of scientific conclusions.

## Tables and Figures

**Table 1.** Summary of Significant Cardiometabolic Divergences (MENA vs. Rest of World)

| Risk Factor | Sex | Age Band | MENA AAPC (%/yr) [Country Range] | Δslope vs. ROW | p-value | FDR q-value |
| --- | --- | --- | --- | --- | --- | --- |
| **Obesity** | Men | 20–29 | +3.49% [+1.60, +5.21] | +0.13 pp/yr | 0.009 | 0.048 |
| **Obesity** | Men | 30–39 | +2.65% [+1.29, +4.24] | +0.11 pp/yr | 0.043 | 0.101 |
| **Diabetes** | Men | 30–39 | +1.90% [–0.76, +4.38] | +0.07 pp/yr | 0.014 | <0.05* |
| **Hypertension** | Women | 30–39 | –0.58% [–3.40, +1.06] | –0.15 pp/yr | 0.001 | <0.05* |
| **Non-HDL cholesterol** | Women | 20–29 | –0.36% [–0.92, +0.18] | –0.0079 mmol/L/yr | 0.017 | <0.05* |
| **Non-HDL cholesterol** | Women | 30–39 | –0.38% [–0.99, +0.19] | –0.0091 mmol/L/yr | 0.017 | <0.05* |

AAPC = Average Annual Percent Change; the bracketed range is the minimum-to-maximum AAPC observed across the 16 MENA countries, not a 95% confidence interval. ROW = Rest of World. pp = percentage points. FDR = False Discovery Rate (Benjamini-Hochberg).
\*The source protocol reports that 5 of these 6 findings survive Benjamini-Hochberg correction at $q < 0.05$ , and states the exact q-value for only two of the six cells (obesity, men 20–29: $q = 0.048$ ; obesity, men 30–39: $q = 0.101$ , the one finding that does *not* survive correction). The precise q-values for the remaining four cells (diabetes men 30–39; hypertension women 30–39; non-HDL cholesterol women 20–29 and 30–39) are not individually reported in the source data — only that they fall under $q < 0.05$ collectively. They are shown here as “<0.05” rather than as a specific numeric value, to avoid stating a precision the source does not provide.

**Figure 1.**
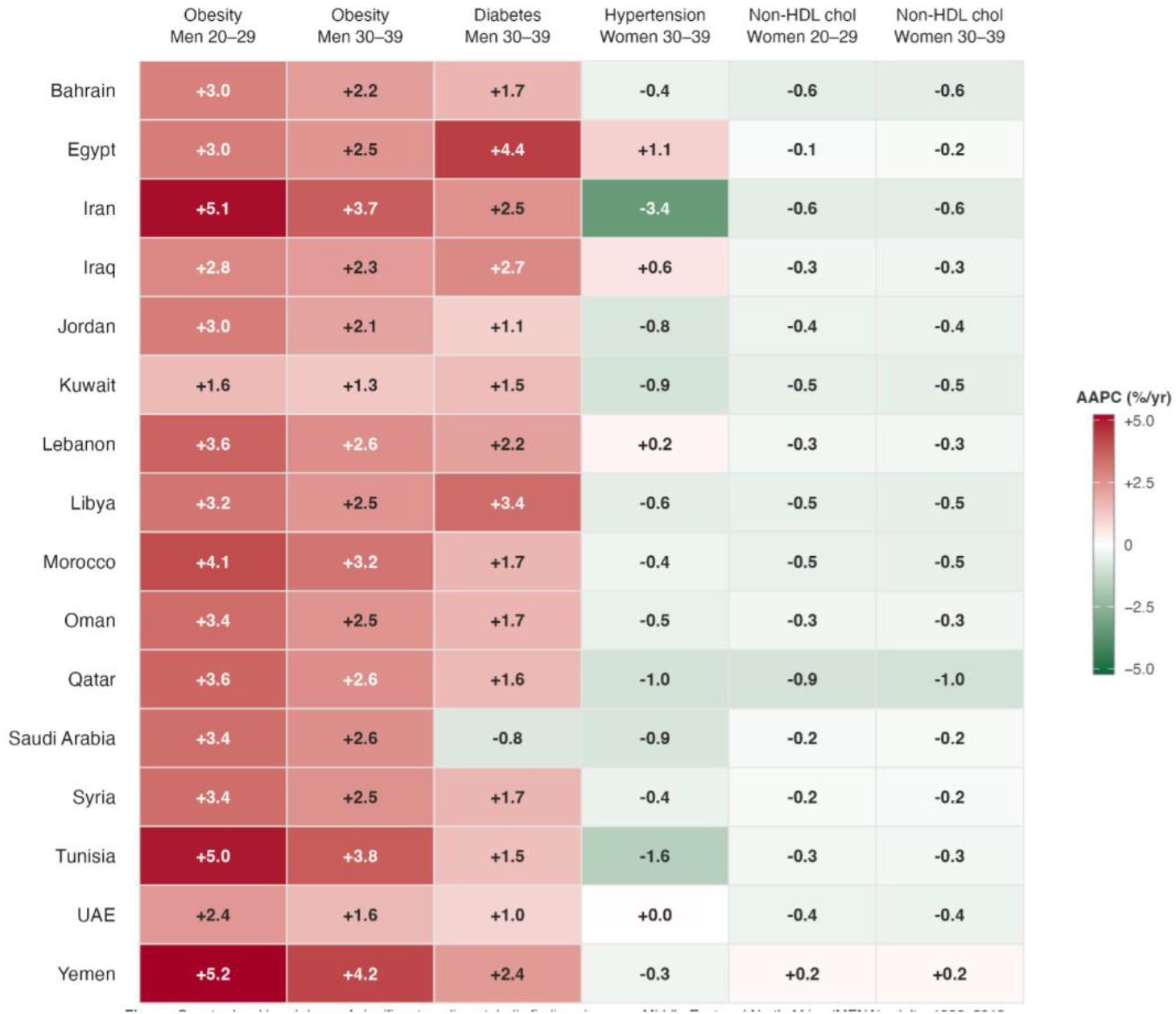
Country-level AAPC heatmap for the six significant cardiometabolic divergences, 16 MENA countries, 1990–2018. Country-level breakdown of the six significant cardiometabolic findings in young Middle East and North Africa (MENA) adults, 1990–2018. Each cell shows the per-country average annual percent change (AAPC) in the named risk factor, for the indicated sex and age band, from joinpoint regression of the annual estimated prevalence (or, for non-HDL cholesterol, mean concentration in mmol/L) on calendar year. The six findings displayed are those for which the MENA-pooled trajectory differed significantly from the population-weighted trajectory of the 184 non-MENA “rest-of-world” (ROW) countries in linear mixed-effects models (year × region interaction, p < 0.05; country-level random intercept and slope). Input data are NCD-RisC country-level Bayesian hierarchical model estimates pooling individual-level data from population-representative surveys (ncdrisc.org); modeled-estimate uncertainty is not propagated into the AAPCs shown here. Hypertension estimates are restricted to adults aged 30 years and older; all other risk factors are shown for both the 20–29 and 30–39 age bands. All analyses were conducted in R 4.5.

